# A signature of platelet reactivity in CBC scattergrams reveals genetic predictors of thrombotic disease risk

**DOI:** 10.1101/2023.07.26.23293204

**Authors:** Hippolyte Verdier, Patrick Thomas, Joana Batista, Carly Kempster, Harriet McKinney, Nicholas Gleadall, John Danesh, Andrew Mumford, Johan Heemskerk, Willem H. Ouwehand, Kate Downes, William J. Astle, Ernest Turro

## Abstract

Genetic studies of platelet reactivity (PR) phenotypes may identify novel antiplatelet drug targets. However, discoveries have been limited by small sample sizes (*n*<5,000) due to the complexity of measuring PR. We trained a model to predict PR from complete blood count (CBC) scattergrams. A GWAS of this phenotype in 29,806 blood donors identified 21 distinct associations implicating 20 genes, of which six have been identified previously. The effect size estimates were significantly correlated with estimates from a study of flow-cytometry measured PR and a study of a phenotype of *in vitro* thrombus formation. A genetic score of PR built from the 21 variants was associated with myocardial infarction and pulmonary embolism. Mendelian randomisation analyses showed PR to be causally associated with the risks of coronary artery disease, stroke and venous thromboembolism. Our approach provides a blueprint for employing phenotype imputation to study the determinants of hard-to-measure but biologically important haematological traits.

**Key points:**

- Platelet reactivity can be predicted from scattergrams generated by haematology analysers of a type in widespread clinical use.
- Genetic analysis of *predicted* platelet reactivity reveals associations with the risks of thrombotic diseases, including stroke.

## Introduction

Platelets are small anucleate blood cells that contribute to physiological clot formation, ensuring haemostasis and healing after vascular injury. However, they also contribute to pathological thrombosis, which underlies venous thromboembolic disease, acute coronary syndrome and ischaemic stroke, including microinfarcts, a leading cause of dementia^1^. Platelets in circulation activate in response to stimulation by agonists such as collagen (which is exposed by injury), adenosine diphosphate (ADP) and thromboxane A2 (which are released by other activated platelets), and thrombin (which is generated by the coagulation cascade). Pathological platelet activation can be caused by atherosclerotic plaque rupture, which exposes collagen and also releases tissue factor, which triggers the coagulation cascade. Antiplatelet therapies are the leading pharmaceutical strategy for primary and secondary prevention of pathological thrombosis and act by reducing platelet reactivity (PR) through blockade of specific activation pathways. For example, clopidogrel inhibits the ADP receptors on the surface of platelets, while aspirin prevents the production of thromboxane A2. The sensitivity of platelets to stimulation is finely balanced. People with inherited platelet disorders that impair PR have a high risk of bleeding^2^. Atherosclerotic patients treated by percutaneous coronary intervention and anti-platelet drugs have a greater risk of bleeding if they have lower PR and a greater risk of forming thrombi that occlude blood vessels, causing heart attacks or strokes, if they have a higher PR^3^. An improved understanding of the mechanisms that govern activation pathways in platelets could unveil novel drug targets with improved pharmacological safety profiles and potentially offer a means to stratify patients better, to maximise the safety and efficacy of existing therapies^4^.

PR is typically characterised using light transmission aggregometry (LTA) to measure the aggregation response of platelets to stimulation by agonists. Alternatively, PR to stimulation by an agonist can be measured by flow cytometry (FC), using surface markers of activation, such as P-selectin, which is externalised during activation, or the binding of fibrinogen to surface receptors following their conformational change during platelet activation. Estimates of the heritabilities of PR phenotypes typically range from 30% to 60%^5–8^. A comparison of two recent studies — one using FC^9^ and the other using LTA^10^ — demonstrates that FC provides a more heritable measure of PR than LTA: evidence of similar strength was obtained for an association between a variant in *GRK5* and PR to thrombin in both studies, but the study using FC relied on half the sample size of the study using LTA. Both LTA and FC approaches are time consuming, difficult to standardise, and require the processing of fresh citrated blood samples soon after venepuncture. This has limited the sample sizes of previous genetic meta-analyses^9–15^ (**Supplemental Table 1**).

To address the limitations of PR and LTA, we explored the possibility of imputing PR phenotypes from measurements made by an alternative technology in widespread clinical use. Sysmex XN haematology analyzers are sophisticated, high-throughput, clinically standardised instruments containing a miniaturised flow cytometer, a device to measure cellular impedance and a spectrophotometer. The primary function of a Sysmex analyser is to generate a complete blood count (CBC) from the data measured by these instruments. A CBC is a standard clinical report that summarises cellular and biochemical properties of the blood, including cell concentrations, cell volume distributions and the concentration of haemoglobin. We hypothesised that the cell-level measurements of platelets generated by the internal flow cytometer of a Sysmex instrument carry information that could be exploited to study PR in large genotyped cohorts. To explore this, we designed a study encompassing three cohorts for which it was possible to access or generate data of different types (**Figure 1a**). Firstly, we generated FC (**Figure 1b**) and Sysmex (**Figure 1c**) data on 533 participants in the Cambridge Platelet Function Cohort (PFC). This allowed us to train models to predict PR to various agonists from cell-level measurements of *unagonised* platelets generated by the Sysmex instrument.

**Figure 1.**
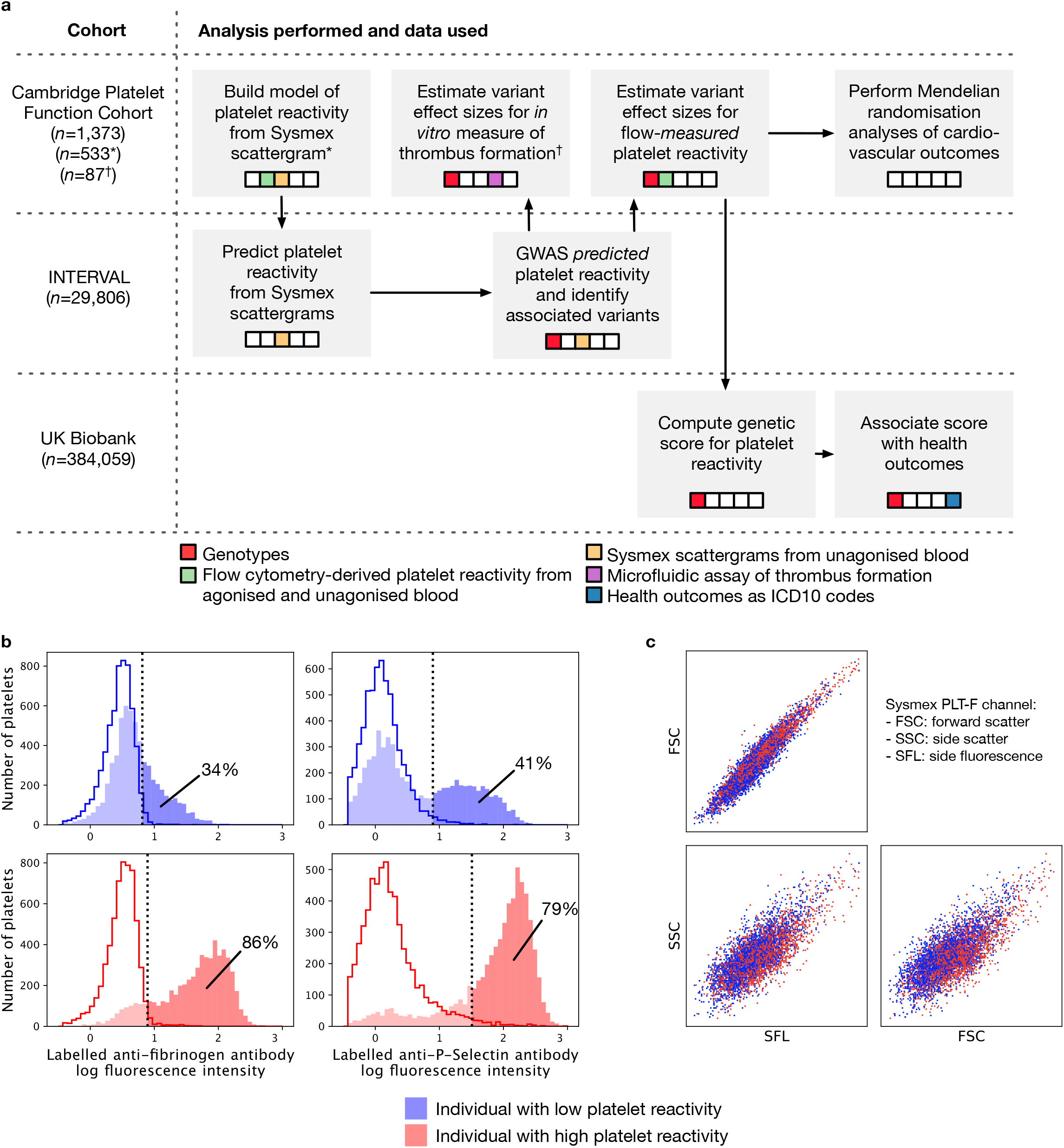
Overview of the study design and data types. **a**, schematic of the analyses; grey blocks represent distinct analyses, which are grouped by row according to the dataset to which they were applied; the arrows illustrate the flow of information between analyses; the colourings of the embedded strips indicate the nature of the primary data used for each analysis. **b**, examples of histograms of log fluorescence intensity from the platelet FC measurements used to quantify PR; the left and right hand panels show measurements of fluorescence generated by an anti-human fibrinogen polyclonal antibody that labels bound fibrinogen and an anti-P-selectin antibody, respectively; the upper and lower panels correspond to an individual with a high (red) and an individual with a low (blue) PR, respectively; the solid histograms represent the distributions of log intensity measurements from activated platelets (using the agonist ADP, in this example) incubated with labelled fibrinogen or a conjugated anti-P-selectin antibody while the open histograms represent negative controls (see **Methods**); the dotted lines correspond to the positivity thresholds (i.e., to the 98% quantiles of the distributions illustrated by the open histograms); the percentages of agonised platelets that exceed the positivity thresholds are shown. **c**, Composite of the two CBC scattergrams measured from blood taken from the two individuals of panel **b**, which are distinguished by colour.

Secondly, we applied the trained models to predict PR to ADP (only PR to ADP could be predicted with useful accuracy) from Sysmex data in 29,806 genotyped participants in the INTERVAL cohort of blood donors^16^. Thirdly, we performed a GWAS of the predicted PR (PPR) phenotype in INTERVAL. Fourthly, we estimated the effects of the variants identified by the GWAS on the means of (i) FC-derived PR phenotypes, and (ii) a phenotype of *in vitro* thrombus formation. Fifthly, we built a genetic score using the variants identified by the PPR GWAS. Sixthly, we computed the genetic score for 384,059 British ancestry participants in the UK Biobank study and tested for associations between the score and 524 health outcomes. Finally, we conducted Mendelian randomisation analyses using large GWAS summary statistics to test for association between PR and the risks of coronary artery disease (CAD), stroke and venous thromboembolism (VTE).

## Methods

A comprehensive description of the methods is given in **Supplemental Methods**. A summary is provided below.

### Measuring FC-derived PR

Platelet activation in response to different agonists was measured using P-selectin and fibrinogen surface markers by whole blood FC according to a previously published protocol^14,17^.

### Isolating platelets in Sysmex XN scattergrams

The PLT-F channel of the Sysmex XN instrument is designed to measure properties of platelets. The channel generates a three dimensional FC scattergram, each point of which corresponds to a cell. We developed a gating procedure to identify platelets (**Supplemental Table 2**) and validated it by comparing the number of cells lying inside the gates with the corresponding PLT value generated by the instrument in each sample (**Supplemental Figure 1**).

### Adjusting for experimental variability in PLT-F scattergrams

We adjusted for between-instrument and time-dependent technical variation in the INTERVAL PLT-F scattergrams using a breakpoint detection algorithm to partition the time series. We adjusted the scattergrams within each class of the partition by applying an affine transformation chosen to match the mean and covariance structure of the scattergram data aggregated within the class to those of the aggregated scattergram data from the PFC.

### Extracting features from PLT-F scattergrams

We developed a procedure based on principal components analysis to extract 15 features capturing inter-individual variation (including measures of central tendency, variation and covariation) from PLT-F scattergrams.

### Building a predictive model of FC-derived PR from Sysmex PLT-F scattergrams

We fitted a Lasso regression^18,19^ model to predict PR in response to a given agonist from the 15 scattergram-derived features and five standard CBC platelet traits (20 features in total)^18,19^ using data from 533 participants in the PFC. We optimised the penalty parameter of the Lasso to maximise the mean (over repetitions and folds) *R*^*2*^ of the regression in held-out data. In the case of ADP, the optimal value of the penalty parameter yielded a mean prediction *R*^2^=0.26 (**Supplemental Figure 2**). Finally, we fitted a linear regression model of the ADP response on the 14 covariates with non-zero coefficients in the cross-validation, using the entire dataset.

### Genome-wide association study of PPR in INTERVAL

We predicted PR in response to ADP in the European participants in the INTERVAL cohort using the model fitted to the PFC data and performed a genome-wide association study of the predicted phenotype using a standard approach. We tested variants with MAF>1% and adopted a significance threshold of *P*<10^−8^.

### Imputation of genotypes in the PFC

We imputed the genotypes in the PFC from a combined 1000 Genomes Phase 3-UK10K whole genome sequencing panel, as described in Downes et al. (ref.^14^).

### Regression of FC-derived phenotypes on variant imputed allele counts

We regressed the four FC-derived PR phenotypes on the imputed allele counts of each of the 21 PPR-associated variants (identified in INTERVAL) using data from 1,373 participants in the PFC^14^. Where necessary, we used the LDproxy tool^20^ to identify the most strongly correlated alternative variant in the reference panel of British in England and Scotland that had been typed in the PFC.

### Replication in Keramarti et al. LTA study

We downloaded the publicly available summary statistics from the LTA study of Keramati et al.^15^, which are restricted to the set of variants with association test *P*-values < 3×10^−4^. We identified the variant in summary statistics exhibiting the strongest LD (*r*^2^ in INTERVAL Europeans) with each of the 21 PPR associated variants and the phenotype exhibiting the smallest *P*-value for association with that variant.

### Regression of an *in vitro* thrombus formation phenotype on variant imputed allele counts

A 48-dimensional *in vitro* thrombus formation phenotype was measured on 87 genotyped participants in the PFC^21^. After standardising each dimension to have a mean of zero and a variance of one, we performed a principal component analysis. We regressed the leading principal component on the imputed variant allele counts corresponding to the 21 PPR-associated variants identified in INTERVAL. We compared the *P*-values and effect sizes identified in the genetic analysis of PPR in INTERVAL and the genetic analysis of the *in vitro* formation phenotype in the PFC. We used proxies for two PPR-associated variants (rs3819288 and rs59001897) as described above.

### Building a genetic score of general PR

For each of the variants identified by the INTERVAL GWAS (or the corresponding proxy, see above), we obtained the vector of previously published PFC effect sizes with respect to FC-measured PR in response to four agonists^14^. We sought to calibrate the effect sizes of the genetic variants, to put them on a scale measuring a general propensity of platelets to activate (a latent, agonist-independent form of PR). Assuming that each causal variant is involved in one of the four measured activation pathways, we linked each variant to the agonist yielding the smallest *P*-value of association in the PFC and assigned it to that pathway. We then applied a standardisation procedure to calibrate the effect sizes corresponding to each agonist. Finally, we computed a polygenic score of general PR as the sum of the linear combinations of genotypes and effect sizes within each of the four agonists.

### Survival analysis in UK Biobank

We performed Cox regression analyses in UK Biobank to test for associations between the genetic score of PR and 524 health outcomes (ICD10 codes) derived from electronic health records, adjusting for several covariates known to play a role in cardiovascular diseases and five standard CBC traits.

### Mendelian randomisation analyses

We performed two-sample Mendelian randomisation analyses to estimate the causal effect of general PR (variant effect sizes calibrated as above) on the log odds of disease events for CAD^22^, stroke^23^ and VTE^24^. We excluded rs61751937 because evidence in the literature suggests variation in *SVEP1* expression may be a risk factor for atherosclerosis^25^. We selected the ten remaining variants in **Table 1** with a *P*-value of association (with a PR phenotype) less than 0.05 in the PFC as primary instruments. We meta-analysed the instrument specific ratio estimates using the standard inverse variance weighted (IVW) fixed effects estimator (**Figure 4e–g**). We then performed a series of concordancy analyses using the robust MR Egger, IVW random effects, weighted median and weighted mode estimators^26–28^.

**Table 1.** GWAS of PPR and genetic score of general PR. The first ten columns show the results of the GWAS of a PR phenotype predicted from Sysmex scattergrams in INTERVAL. The coordinates are given with respect to genome build GRCh37. As statistics summarising the associations in the PFC between PR phenotypes and the variants with rsIDs marked with a ‘ □’ were not available, we identified suitable proxies in the PFC to build a genetic score (given in **Supplemental Table 3**). The subsequent columns include (i) a comment on each gene mapped to the associated SNPs, (ii) a list of CBC trait associations identified previously by GWAS^59^ and (iii) the effect size and *P*-value corresponding to the PR trait with the smallest *P*-value for association in the PFC and the agonist corresponding to that trait. The bold gene names indicate loci previously associated with a PR phenotype by GWAS (at *P*<5×10^−8^) or gene-based (at *P*<10^−5^) analyses (marked with an asterisk). The underlined gene names indicate associations with evidence for replication in partial summary statistics from LTA studies (at *P*<3×10^−4^)^10,15^.

| Chr | Position | rsID | Ref | Alt | MAF | P<sub>INTERVAL</sub> | β<sub>INTERVAL</sub> | SE<sub>INTERVAL</sub> | Gene | Comments | CBC associations | β<sub>score</sub> | P<sub>score</sub> | Phenotype for β<sub>score</sub> |
| --- | --- | --- | --- | --- | --- | --- | --- | --- | --- | --- | --- | --- | --- | --- |
| 1 | 156869047 | rs12566888 | G | T | 0.10 | 5.00x10<sup>-24</sup> | -0.12 | 0.012 | <u>PEAR1</u> | Variant is associated with ADP and epinephrine induced platelet aggregation<sup>11,12</sup>. Platelet receptor that signals upon platelet-platelet contact in response to and independently of the activation response<sup>33</sup>. | PLT, MPV, PDW | -0.99 | 3.20x10<sup>-8</sup> | ADP |
| 1 | 199010721 | rs1434282 | C | T | 0.27 | 1.90x10<sup>-17</sup> | -0.07 | 0.008 | <u>PTPRC</u> | Protein regulates GP6-mediated signalling during platelet activation via SRC-family kinase<sup>37</sup>. | PLT, MPV, PCT | -0.12 | 4.50x10<sup>-3</sup> | CRP-XL |
| 1 | 247712303 | rs41315846 | T | C | 0.47 | 1.40x10<sup>-11</sup> | -0.05 | 0.007 | <u>GCSAML</u> | Protein identified as a mediator of platelet activation downstream of cAMP/protein kinase A signalling using protein-protein interaction analysis<sup>38</sup>. | PLT, MPV, PDW, PCT | -0.17 | 1.10x10<sup>-3</sup> | PAR1 |
| 2 | 224874874 | rs13412535 | G | A | 0.24 | 2.20x10<sup>-10</sup> | -0.06 | 0.009 | <u>SERPINE2</u> | Protein is a serine protease inhibitor with activity against the potent platelet activator thrombin<sup>39</sup>. | MPV | 0.07 | 1.3x10<sup>-1</sup> | CRP-XL |
| 2 | 241510903 | rs78909033 | G | A | 0.13 | 2.50x10<sup>-11</sup> | -0.07 | 0.010 | <u>RNPEPL1</u> |  | PLT, MPV, PDW, PCT | 0.10 | 6.60x10<sup>-2</sup> | CRP-XL |
| 3 | 56849749 | rs1354034 | T | C | 0.40 | 1.20x10<sup>-24</sup> | -0.07 | 0.007 | <u>ARHGEF3</u> | Variant is associated with ADP-induced aggregation<sup>14</sup>. Guanine nucleotide exchanger for Rho A implicated in platelet production and ADP-mediated platelet activation<sup>31</sup>. | PLT, MPV, PDW, PCT | -0.64 | 4.20x10<sup>-10</sup> | ADP |
| 3 | 124340093 | rs13067286 | G | A | 0.48 | 2.20x10<sup>-11</sup> | 0.05 | 0.007 | <u>KALRN</u> | Protein is a Rho-GTPase activator activated by ADP receptor signalling in platelets<sup>40</sup>. Another variant in *KALRN* (rs3772800) is associated with the risk of myocardial infarction<sup>41</sup>. | PLT, MPV, PDW | -0.14 | 4.90x10<sup>-3</sup> | PAR1 |
| 3 | 124366890 | rs76445378 | C | T | 0.02 | 5.80x10<sup>-09</sup> | -0.17 | 0.029 | <u>KALRN</u> |  | PLT, MPV, PDW | -0.41 | 8.40x10<sup>-2</sup> | PAR1 |
| 5 | 122088890 | rs922140 | A | G | 0.40 | 9.40x10<sup>-21</sup> | 0.07 | 0.007 | <u>SNX2</u> |  | MPV, PCT | -0.08 | 1.30x10<sup>-1</sup> | PAR1 |
| 6 | 31322694 | rs3819288† | T | C | 0.10 | 1.20x10<sup>-12</sup> | -0.08 | 0.012 | <u>HLA-B</u> | Antigen-presenting major histocompatibility complex class I molecule that mediates alloimmune clearance of circulating platelets<sup>42</sup>. Other variants in region are associated with unstable angina pectoris<sup>43</sup>, myocardial infarction, use of antithrombotic agents<sup>44</sup> and rheumatoid arthritis<sup>45</sup> amongst many other phenotypes. | PLT, MPV | 0.19 | 1.80x10<sup>-3</sup> | PAR4 |
| 9 | 99234329 | rs55665228 | C | T | 0.19 | 4.00x10<sup>-11</sup> | -0.06 | 0.009 | <u>HABP4</u> |  | PLT, MPV, PDW, PCT | -0.35 | 1.20x10<sup>-2</sup> | ADP |
| 9 | 113312231 | rs61751937 | G | C | 0.03 | 9.20x10<sup>-16</sup> | 0.17 | 0.021 | <u>SVEP1*</u> | Gene recently associated with ADP-induced platelet aggregation using a gene-based approach<sup>15</sup>. Extracellular matrix protein that interacts and activates with PEAR1 in platelets<sup>46</sup>. | PLT, MPV, PDW | 1.45 | 8.20x10<sup>-6</sup> | ADP |
| 10 | 121010256 | rs10886430 | A | G | 0.14 | 2.30x10<sup>-10</sup> | 0.07 | 0.011 | <u>GRK5</u> | Variant is associated with thrombin-induced platelet aggregation and VTE<sup>10,14</sup>. The protein is a serine/threonine kinase GPCR regulator that modulates thrombin-mediated platelet activation<sup>14</sup>. | PLT, MPV, PDW | 0.90 | 2.80x10<sup>-40</sup> | PAR1 |
| 11 | 10711817 | rs7123827 | A | C | 0.50 | 7.20x10<sup>-10</sup> | 0.04 | 0.007 | <u>IRAG1</u> | Other variants in region are associated with ADP and epinephrine-induced platelet aggregation<sup>11,12</sup>. Inositol 1,45-triphosphate receptor regulator during nitric oxide/cyclic GMP modulation of platelet reactivity<sup>47</sup>. | PLT, MPV | 0.05 | 1.70x10<sup>-1</sup> | CRP-XL |
| 12 | 122216910 | rs11553699 | A | G | 0.15 | 4.20x10<sup>-35</sup> | 0.13 | 0.011 | *RHOF* | Variant is associated with serum levels of heparin-binding EGF<sup>48</sup>. Small GTPase actin regulator that mediates platelet filopodia formation<sup>49</sup>. | PLT, MPV, PDW, PCT | 0.06 | 3.20x10<sup>-1</sup> | CRP-XL |
| 14 | 70653758 | rs61978213 | G | A | 0.05 | 2.40x10<sup>-12</sup> | 0.12 | 0.018 | *SLC8A3* | Protein is a sodium/calcium exchanger involved in platelet calcium hemostasis<sup>50</sup>. Another variant near gene (rs55784307) is associated with peripheral arterial disease<sup>51,52</sup>. |  | 0.20 | 5.90x10<sup>-2</sup> | PAR4 |
| 15 | 65160392 | rs59001897† | T | A | 0.18 | 6.40x10<sup>-09</sup> | 0.05 | 0.010 | *PLEKHO2* | Another variant near *PLEKHO2* (rs832890) is associated with pulse pressure<sup>53</sup> | PLT, MPV, PDW, PCT | 0.15 | 2.80x10<sup>-1</sup> | ADP |
| 16 | 9052989 | rs8057254 | T | A | 0.19 | 2.80x10<sup>-09</sup> | 0.05 | 0.009 | *USP7* | Inhibition of USP7 blocks collagen-stimulated aggregation<sup>54</sup>. | PLT, MPV, PDW | 0.10 | 3.40x10<sup>-2</sup> | CRP-XL |
| 16 | 81870969 | rs12445050 | C | T | 0.14 | 5.20x10<sup>-25</sup> | 0.11 | 0.010 | *PLCG2* | Variant associated with VTE<sup>55</sup> and CAD<sup>43</sup>. Protein is a member of the phospholipase C family and mediates GP6 and αIIbβ3-mediated platelet activation<sup>56</sup>. | PLT, MPV, PDW | 0.09 | 1.00x10<sup>-1</sup> | CRP-XL |
| 17 | 3819002 | rs11078475 | T | C | 0.47 | 2.50x10<sup>-10</sup> | 0.05 | 0.007 | *P2RX1* | Gene is overexpressed in reticulated platelets<sup>57</sup>. Protein is a platelet ATP receptor contributing to platelet granule release<sup>58</sup>. | PLT, MPV | 0.16 | 1.10x10<sup>-1</sup> | ADP |
| 19 | 55538980 | rs1654425 | T | C | 0.17 | 3.70x10<sup>-22</sup> | 0.09 | 0.009 | *GP6* | A variant in strong LD (rs1671152) is associated with collagen-induced platelet aggregation mediated by surface receptor encoded by *GP6*<sup>12</sup>. Protein is a platelet receptor for the potent activating agonist collagen. | PLT, MPV, PDW | 0.89 | 5.40x10<sup>-100</sup> | CRP-XL |

## Results

We attempted to predict four FC-derived phenotypes measuring PR in response to each of ADP, a synthetic cross-linked collagen-related peptide (CRP-XL^29^), and two peptides targeting the thrombin receptors PAR-1 and PAR-4 respectively from haematology analyser data. Linear regressions of each of the four phenotypes measuring PR to agonists on the five standard platelet CBC traits (platelet count (PLT), mean platelet volume (MPV), platelet crit (PCT), platelet distribution width (PDW) and immature platelet fraction (IPF)) showed poor predictive performance (all *R*^*2*^≤0.10). Consequently, we applied a gating procedure to select platelets from the PLT-F channel scattergrams and extracted 15 quantitative summary features from each of the resulting sub-scattergrams (**Methods, Supplemental Table 2**). We fitted Lasso regressions to predict each of four FC-derived phenotypes measuring PR to agonists from the 15 features and 5 platelet CBC traits, using data from 533 individuals in the PFC. We used 100 repetitions of five-fold cross-validation to tune the Lasso penalty parameters. While PR in response to each of CRP-XL, PAR-1 and PAR-4 could not be predicted usefully (mean *R*^*2*^ in held-out data <0.05, discussed below), PR in response to ADP could be predicted with mean *R*^*2*^=0.26. Consequently, we investigated whether a GWAS of this phenotype using tens of thousands of individuals could identify genetic variation related to PR.

We fitted the tuned Lasso regression for PR to ADP without holding data out. Using this fitted model, we predicted PR to ADP from Sysmex PLT-F scattergram data generated on 29,806 participants in the INTERVAL cohort. We performed univariable tests for additive allelic association between the PPR phenotype and genotypes at 10,013,294 imputed genetic variants. Using stepwise regression we identified a parsimonious subset of 21 variants explaining the significant (at *P*<10^−8^) genetic associations with PPR and determined the nearest protein coding gene to each variant (**Figure 2, Table 1**).

**Figure 2.**
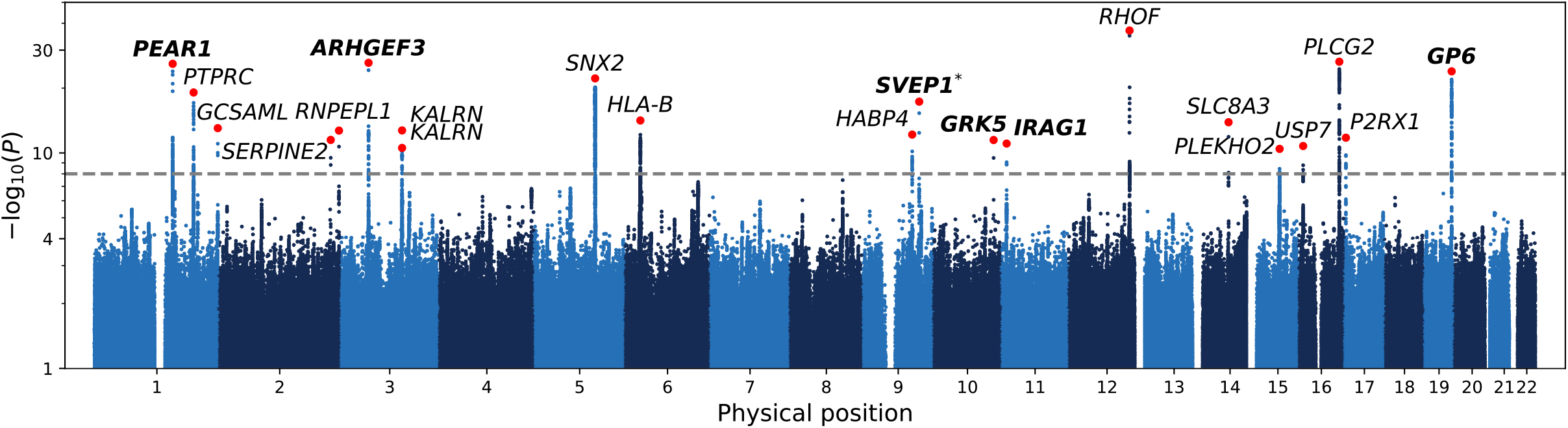
Genome-wide associations with PPR. A Manhattan plot showing the *P*-values of linear mixed model tests for association between genetic variants and PR predicted from CBC scattergrams in INTERVAL. Each dot corresponds to a genetic variant in the Haplotype Reference Consortium r1.1 reference panel. The position on the *x*-axis indicates the physical position of the variant; the position on the *y*-axis indicates -log10 of the *P*-value (on the log scale) corresponding to the χ^2^ BOLT-LMM statistic. Only variants with an imputation INFO score greater than 0.4 and a *P*-value less than 0.1 are shown. The horizontal dashed line corresponds to the genome-wide significance threshold (α*=*10^−8^). The red dots correspond to the variants showing the strongest evidence for association in those loci containing significantly associated variants. The gene names indicate the protein-coding gene that is nearest to each of these associations. The genes in bold mark loci previously associated with a PR phenotype by GWAS (at *P*<5×10^−8^) or gene-based (at *P*<10^−5^) analyses (marked with an asterisk).

Of the 20 genes identified, *ARHGEF3, GP6, GRK5, IRAG1* and *PEAR1* have been implicated as mediators of variation in PR previously by GWAS (**Table 1**). Although no genome-wide significant single-variant association implicating *SVEP1* in the mediation of PR had been reported prior to the present study (rs61751937, *P*=9.20×10^−16^), a gene-based genetic association between *SVEP1* and PR has recently been reported with a border-line *P*-value (*P*=2.6×10^−6^, □=2.82×10^−6^). However, this test relied principally on evidence from a single variant (rs61751937, univariable *P*=5.84×10^−6^, the same variant identified in the present study)^15^.

*ARHGEF3* encodes an MK-expressed RhoGEF that has been previously associated with PR^14,30,31^. *GP6* encodes one of the two major collagen receptors on the surface of platelets. *GRK5* is a G protein-coupled receptor kinase that regulates thrombin signalling, possibly by phosphorylating the receptors PAR-1 and PAR-4, leading to their internalisation and destruction^10,14^. *IRAG1* plays a role in the inhibition of platelet aggregation and *in vivo* thrombosis in mice^32^. *PEAR1* encodes an endothelial aggregation receptor that signals secondarily to α_IIb_β_3_-mediated contact between platelets^33^. *SVEP1* encodes a protein that may mediate variation in PR through cell-cell adhesion, cell differentiation or mechanisms in bone marrow niches^15,34^. All but three of the remaining 14 genes tagged by variants associated with PPR in INTERVAL have plausible roles in biological processes underlying platelet activation (**Table 1**).

Of the 6 genes previously implicated in the variation of PR phenotypes by genetic association analyses, *PEAR1, ARHGEF3, SVEP1* and *IRAG1* mediate associations with PR to ADP; *PEAR1* also mediates PR to epinephrine; *GRK5* mediates PR to thrombin; and *GP6* mediates PR to collagen. The GWAS of PPR (to ADP) therefore had power to identify genes playing a role in multiple PR pathways, suggesting that the predictive signature in the Sysmex scattergrams captures biological variation downstream of the convergence point of the activation pathways initiated by these different agonists.

To strengthen the evidence that the PPR phenotype derived from Sysmex scattergrams can be a useful proxy for identifying associations with PR in general, we tested the 21 variants associated with PPR for association with each phenotype measuring an agonist-induced PR response in the PFC. We regressed the four FC-measured PR phenotypes (measuring responses to ADP, CRP-XL, PAR1 and PAR4 respectively) on the imputed allele count of each variant (**Supplemental Table 3**). The *P*-values for each agonist were skewed towards zero relative to the uniform distribution on (0,1) (**Figure 3a**). When controlling the false discovery rate (FDR, Benjamini–Hochberg procedure) at 0.05, five variants were significantly associated with PR to ADP, three variants with PR to CRP-XL, three variants with PR to PAR1 and three variants with PR to PAR4. The variants tagging four genes previously implicated in variation of PR—*GP6, GRK5, ARHGEF3, PEAR1* and *SVEP1*—exhibited the strongest evidence for association with the PR phenotypes, having minimum *P*-values across the agonists ranging from 5.37×10^−100^ to 8.24×10^−6^. Variants tagging four genes that had not been previously implicated in variation of PR—*GCSAML, HLA-B, PTPRC* and *KALRN*—exhibited minimum *P*-values ranging between 4.91×10^−3^ and 1.05×10^−3^, strongly suggesting that the analysis of PPR can reveal novel mediators of PR. To demonstrate that the PPR associations are enriched for associations with PR phenotypes relative to standard CBC platelet traits, we compared the distribution of minimum (over PR phenotypes) *P*-values between the PPR-associated variants and variants associated with standard CBC traits in INTERVAL^35^. The distribution of *P*-values for the PPR-associated variants was lower (**Supplemental Figure 3**).

**Figure 3.**
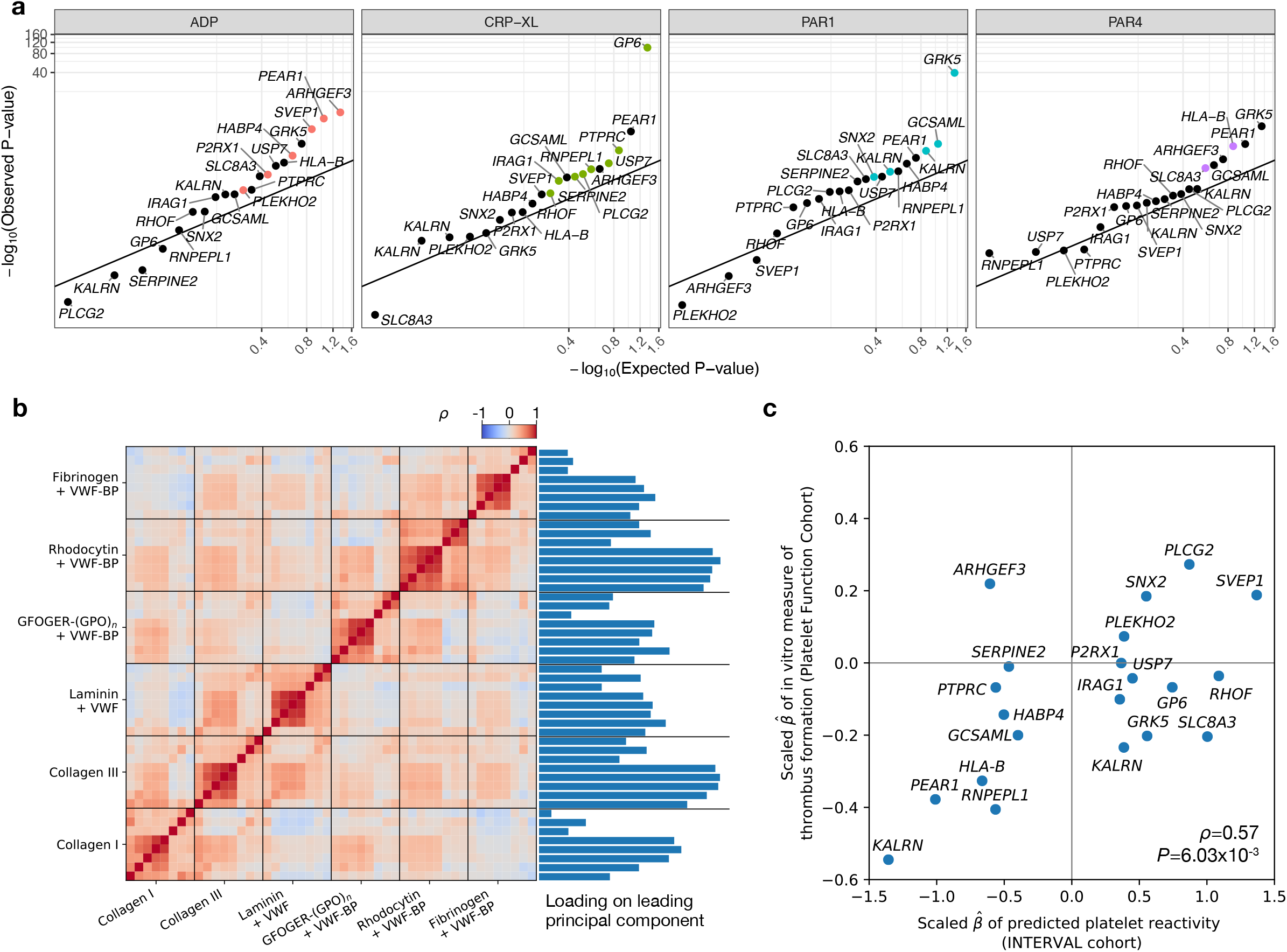
Regression of PR-related phenotypes on PPR-associated variants. **a**, Log_10_ scale Q-Q plot comparing the *P*-values obtained by regressing FC-derived PR to ADP, CRP-XL, PAR1 and PAR4 on the variant imputed allele counts at 21 PPR-associated sites in 1,373 PFC participants with the *P*-values obtained under the null hypothesis of no association. The smallest *P*-value obtained across agonists for each variant is highlighted using an agonist-specific colour. **b**, Heatmap showing the correlation structure among 48 *in vitro* phenotypes of thrombus formation and their relative loadings on the leading principal component of a principal components analysis. **c**, Scatterplot of the scaled effect sizes of the 21 PPR-associated variants with respect to PPR in INTERVAL and the first principal component of the *in vitro* thrombus formation phenotypes in 87 PFC participants. The correlation (ρ) and the *P*-value under the null hypothesis ρ=0 are embedded.

**Figure 4.**
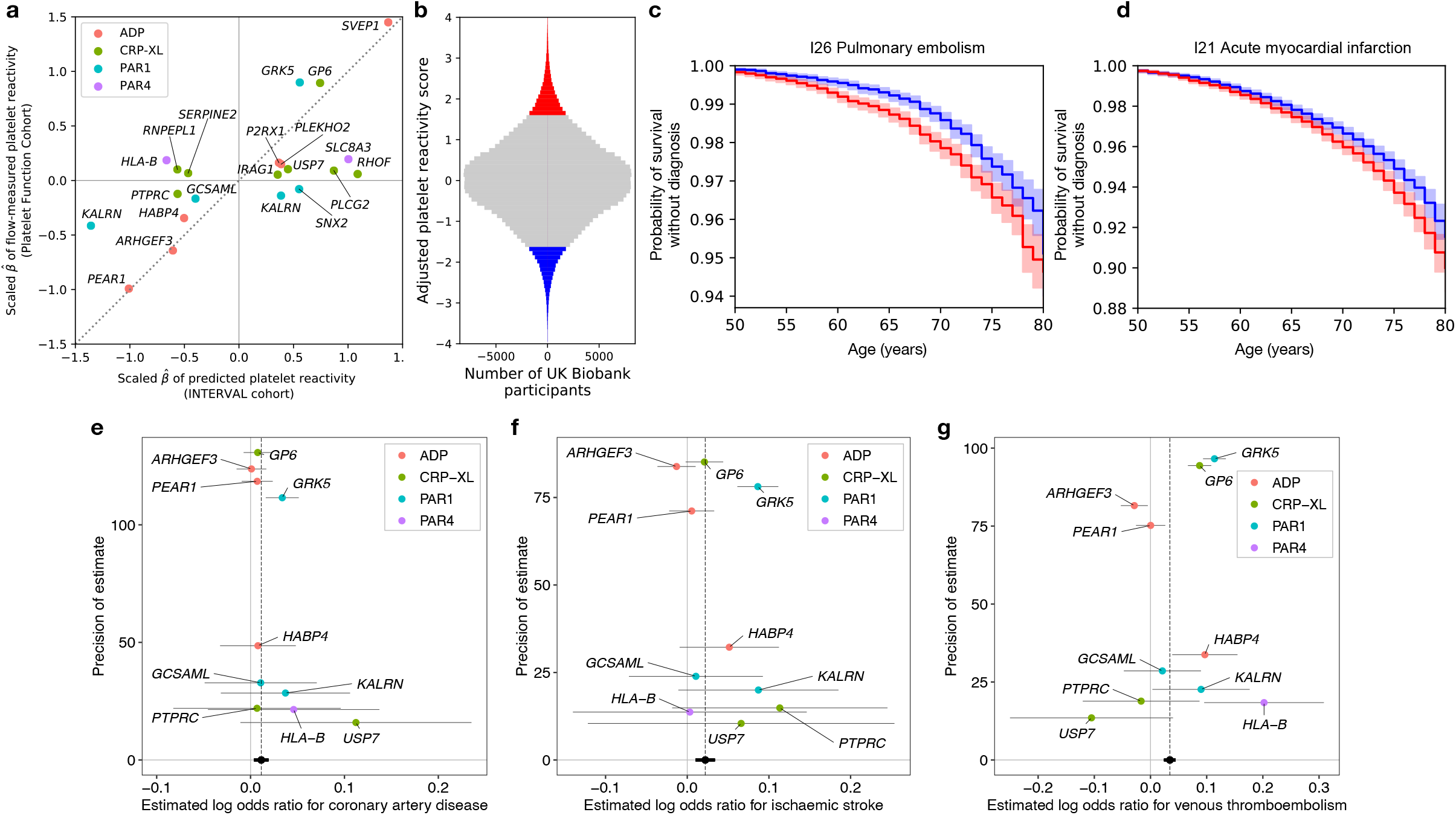
Genetic score of PR and its association with health outcomes. **a**, Scatterplot, each dot of which corresponds to a variant identified by a stepwise multiple regression analysis of genetic associations with PPR in INTERVAL. The *x*-axis shows the estimated additive effect size of each variant on mean PPR in INTERVAL. The *y*-axis shows the estimated additive effect size of each variant on the mean of the agonist-specific PR phenotype with the smallest *P*-value in the PFC. The two sets of effect sizes were scaled in order to render them commensurable, as described in **Methods. b**, Histogram of the genetic score for PR, after adjusting for known risk factors and platelet traits, computed in UK Biobank participants, with participants in the upper and lower vigintiles of the distribution marked in red and blue, respectively. **c**,**d**, Kaplan-Meier plots corresponding to the two vigintile subgroups for the two health outcomes that were significantly associated with the genetic score. Plain lines indicate the estimated survival functions and the shaded areas represent the 90% confidence intervals. **e–g**, Funnel plots showing estimates of the log odds ratio for a disease event per standard deviation increase in PPR. The horizontal line segments represent the corresponding 95% confidence intervals. The black circle and line segment lying on the *x*-axis show the estimate and confidence interval for the log odds ratio derived from an inverse variance weighted fixed effects meta-analysis. Confidence intervals were derived using second order weights. Each point is coloured according to the PR phenotype used to calculate the general PR effect size of the corresponding variant.

We sought to replicate the PPR associations using publicly available GWAS summary statistics of LTA-measured PR phenotypes from Keramati et al.^15^. PPR-associated variants in five loci — *PEAR1, ARHGEF3, GP6, SVEP1*, and *PTPRC* — were in strong LD (*r*^2^>0.8) with a variant exhibiting an LTA association (*P*<3×10^−4^). PPR-associated variants in three loci — *HLA-B* (*r*^*2*^=0.47), *IRAG1* (*r*^*2*^=0.35) and *USP7* (*r*^*2*^=0.07) — were in moderate LD with a variant exhibiting an LTA association (*P* < 3×10^−4^). Finally, two PPR-associated variants (*GCSAML* and *PLCG2*) were within 10kb but not in LD with a variant exhibiting an LTA association (*P*<3×10^−4^), providing supporting evidence that these two genes are mediators of variation in PR. The *GRK5* variant has been associated with LTA phenotypes in a separate study^10^. Therefore, of the 20 genes identified by the PPR GWAS, we found replicative evidence from GWAS of LTA phenotypes for 11 genes.

To assess whether variants identified by the genetic analysis of PPR are associated with the tendency of blood to form thrombi, we analysed previously published measurements made with an *in vitro* assay of thrombus formation performed on fresh blood samples from 87 PFC participants^21^. Briefly, glass coverslips were coated with six microspots, each containing a different platelet agonist. Whole blood was perfused onto the microspots using a parallel-plate flow chamber. Eight variables representing phenotypes related to platelet adhesion, aggregation or activation were measured on each microspot using a fluorescence microscope. To account for the correlation structure between the 48 parameters of thrombus formation, we performed dimensionality reduction by principal components analysis and regressed the leading principal component on the imputed allele count of each variant (**Figure 3b**). Although the sample size was insufficient for any variant to exhibit a statistically significant association (**Supplemental Table 3**), the effect size estimates were significantly correlated with the effect size estimates for PPR from INTERVAL (**Figure 3c**, ρ=0.57, *P*=6.03×10^−3^), providing good evidence that some variants associated with PPR play a role in the formation of thrombi.

To explore whether variation in PR might be a predictor of health outcomes, we built a genetic score of PR using the 21 variants identified by the GWAS of PPR. Although the functions of the genes proximal to the variants associated with PPR imply that most are also associated with PR, we were cautious about relying on effect sizes estimated by the GWAS of PPR to weight the genetic score. PPR is only weakly predictive of the FC-measured PR to ADP (*R*^*2*^=0.26), so in principle the phenotype could have a component of variation depending on biological mechanisms extraneous to PR. We therefore sought to identify an estimate of the effect of each variant on a general propensity of platelets to activate (general PR), unbiased by extraneous variation. We assumed that the effect of each variant on PR is mediated by one of the pathways activated by ADP, CRP-XL, PAR1 and PAR4. We assigned each variant *ad hoc* to the pathway corresponding to the agonist yielding the smallest *P*-value of association in the PFC. We standardised the estimated effect sizes to render them commensurable across PR phenotypes (i.e., FC-measured PR to ADP, CRP-XL, PAR1 and PAR4) and used the standardised estimates to weight the imputed allele counts in the polygenic score (**Methods**). The weights assigned to the 21 variants were only moderately correlated with the effect sizes for association with PPR (**Figure 4a**, *R*^*2*^=0.47), so the genetic score of general PR differs substantially from the score that would be derived from the effect size estimates of the PPR GWAS.

We computed the genetic score of general PR for 384,059 British-ancestry participants in UK Biobank. For each of 524 ICD10 codes recording diagnostic events in at least 1,000 participants, we applied Cox proportional hazards regression to estimate the association between the survival time from birth to the event and the genetic score of PR (**Methods**). To adjust the estimates for variation mediated through known risk factors for cardiovascular disease, we included the following variables as covariates in each regression: sex, tobacco use, total cholesterol level, HDL cholesterol level, systolic blood pressure, C-reactive protein concentration and history of diabetes^36^. To ensure that any identified associations were mediated independently of standard platelet parameters, we also included the four platelet traits measured in UKB (PLT, MPV, PCT and PDW) as covariates. The score was significantly (Bonferroni controlled family-wise error rate of 0.05) associated with two ICD10 codes, both of which record cardiovascular events with an aetiological link to PR: pulmonary embolism (I26) (*P*=5.14×10^−8^) and acute myocardial infarction (I21) (*P*=5.50×10^−6^) (**Table 2**). We compared the survival distributions of the individuals in the upper and lower 5% tails of the score distribution. The time required to achieve a 2% cumulative probability of a pulmonary embolism diagnosis was approximately 3 years longer for the lower tail than the upper tail, while the time required to achieve a 5% cumulative probability of an acute myocardial infarction diagnosis was approximately 2 years longer for the lower tail than the upper tail (**Figure 4b–d**).

**Table 2.** Cox regression association statistics. Significant associations (at a family-wise discovery rate <0.05, equivalent to *p*<1.0×10^−4^) between the genetic score of PR and ICD10-coded health outcomes in 384,059 unrelated British-ancestry participants in UK Biobank. ICD10 sub-terms (i.e., containing a ‘.’) were collapsed to the parent term. Only collapsed terms assigned to at least 1,000 participants were analysed (495 terms in total).

| *P* | ICD10 code | Description | No. of participants with an event | Log hazard ratio |
| --- | --- | --- | --- | --- |
| 5.14×10<sup>-8</sup> | I26 | Pulmonary embolism | 6,942 | 0.07 |
| 5.50×10<sup>-6</sup> | I21 | Acute myocardial infarction | 14,198 | 0.04 |

Next, we sought to validate the association between the genetic component of variability in general PR and cardiovascular outcomes using recently published large case-control GWAS of CAD, stroke and VTE^22–24^. Without access to individual level data, we were unable to compute the genetic score in the study participants, so instead we performed two-sample Mendelian randomisation analyses using the two sample inverse variance weighted (IVW) estimator and complementary robust methods^26^. After removing weak instruments (minimum *P*-value in the PFC >0.05) and the variant in *SVEP1*, which may affect vascular risk through a horizontal pathway^25^, ten variants remained. As at least one of these was assigned to each of the four agonists in the construction of the score, variation in PR mediated through all four pathways contributed to the analyses. Variation in PR was significantly and positively associated with the risks of CAD (*P*=0.019), stroke (*P*=2.58×10^−4^) and VTE (*P*=6.55×10^−11^) (**Figure 4e–g**). None of the estimates of the intercepts of the Egger regression models differed significantly (*P*-value>0.05) from zero, implying an absence of evidence for directional pleiotropy. Point estimates and confidence intervals derived from the complementary robust estimators were broadly consistent with the IVW estimates (**Supplemental Table 4, Supplemental Figure 4**).

## Discussion

Despite its clinical importance, PR is challenging to measure, which has limited GWASs of PR phenotypes to sample sizes of a few thousand participants (**Supplemental Table 2**). Our GWAS of PPR in 29,806 blood donors was able to identify variants known to be associated with PR to various agonists without the need for technically challenging platelet stimulation experiments, by exploiting previously unrecognised information on PR contained in Sysmex XN scattergrams derived from EDTA-treated whole blood. We suspect that the variants can be identified in this way because the blood contains small quantities of ADP, collagen and thrombin, and inter-individual variation of PR to these agonists generates variation downstream of the convergence of the activation pathways, which is reflected in the Sysmex scattergrams. It may be that the scattergram signature reflects variation in the stimulation of platelet surface receptors, stimulated in sufficient numbers to cause morphological changes but insufficient numbers to cause activation. When training from Sysmex data, we were able to predict FC-measured PR to ADP better (*R*^*2*^=0.26) than PR to CRP-XL, PAR1 and PAR4 (all *R*^*2*^ <0.05).

Furthermore, our GWAS of PPR produced much smaller *P*-values for genetic variants known to cause variation in PR to ADP (*PEAR1, ARHGEF3*) than for genetic variants known to cause variation in PR to collagen (*GP6*) or thrombin (*GRK5*) (**Figure 2**), despite the fact that the power to detect the associations by FC was lower for PR to ADP than for PR to collagen or thrombin^14^. We speculate this is because ADP is present in EDTA-treated blood in more potent quantities than collagen or thrombin.

We identified six genes previously found by GWAS or by gene-based association analysis of PR phenotypes, including all three of the genes previously identified in at least two non-overlapping study cohorts: *GP6, GRK5* and *PEAR1* (**Supplemental Tables 1 and 3**). In addition, we identified 14 highly credible candidate genes. For example, one of the candidates, *SERPINE2*, encodes Serpin Family E Member 2, a natural inhibitor of thrombin, a strong platelet activator that binds to protease-activated receptors on the surface of platelets. The 21 SNPs identified by our GWAS were collectively associated with an *in vitro* measure of thrombus formation, supporting the hypothesis that the identified genes mediate biological mechanisms involved in thrombosis.

Detailed laboratory follow up of these mechanisms, beyond the scope of the present study, will be required to determine whether they present viable drug targets.

We sought to identify causal associations between PR phenotypes and health outcomes using genetics. However, because PPR is not a direct measure of PR, the effect sizes computed from the INTERVAL GWAS of PPR were potentially biased as estimates of PR. Consequently, we used estimates of effect sizes for association with PR phenotypes in the PFC to quantify the variation in general PR explained by each of the genetic variants associated with PPR, decoupling detection from estimation. We demonstrated the effectiveness of this approach by showing that a genetic score of general PR predicts health outcomes that are closely linked to platelet function, namely survival without pulmonary embolism and survival without acute myocardial infarction. In addition, two-sample Mendelian randomisation analysis demonstrated an association between variation in PR and the risks of CAD, stroke and VTE. These results represent the first time the causality of PR as a risk factor for cardiovascular events has been demonstrated using genome-wide instrumental analyses. Other difficult-to-measure risk factors with correlates that are easy to measure may benefit from a similar approach.

## Data Availability

Access to de-identified INTERVAL participant data can be sought by emailing the INTERVAL Data Access Committee at. Access to the genotype, FC, Sysmex scattergram and microspotting data from PFC participants can be sought by emailing the corresponding author. The genotype data for the genotyped participants in UK Biobank are available via application at https://www.ukbiobank.ac.uk/enable-your-research/apply-for-access. The GWAS summary statistics for CAD, stroke and VTE are available from https://www.ebi.ac.uk/gwas/studies/GCST90132314, https://www.ebi.ac.uk/gwas/studies/GCST90104539 and https://download.decode.is/form/2022/vte_meta.txt.gz, respectively. The code for the analysis is available at https://github.com/hippover/sysmex2pf.

https://www.ebi.ac.uk/gwas/studies/GCST90132314

https://www.ebi.ac.uk/gwas/studies/GCST90104539

https://download.decode.is/form/2022/vte_meta.txt.gz

## Acknowledgements

This research has been conducted using the UK Biobank Resource under Application Number 13745. We are grateful to the Lowy Foundation USA for supporting this work. We are grateful to Mr Stephen Garner for generating part of the Cambridge PFC FC dataset and for his invaluable advice on the operation of Sysmex haematology analysers. We thank Drs. Jarob Saker and Joachim Linssen of Sysmex Europe and Rob Gillions of UK Biobank for invaluable technical assistance and advice. We gratefully acknowledge the participation of all UK Biobank, NIHR Cambridge BioResource, and INTERVAL volunteers. We thank members of the Cambridge BioResource Scientific Advisory Board and Management Committee for their support of our study and the NIHR Cambridge Biomedical Research Centre for funding (RG64219). Participants in the INTERVAL randomised controlled trial were recruited with the active collaboration of NHS Blood and Transplant England, which has supported field work and other elements of the trial. DNA extraction and genotyping were co-funded by the National Institute for Health and Care Research (NIHR), the NIHR BioResource and the NIHR Cambridge Biomedical Research Centre (BRC-1215-20014). The academic coordinating centre for INTERVAL was supported by core funding from the: NIHR Blood and Transplant Research Unit in Donor Health and Genomics (NIHR BTRU-2014-10024), NIHR Blood and Transplant Research Unit in Donor Health and Behaviour (NIHR203337), UK Medical Research Council (MR/L003120/1), British Heart Foundation (SP/09/002; RG/13/13/30194; RG/18/13/33946) and NIHR Cambridge BRC (BRC-1215-20014; NIHR203312). The views expressed are those of the authors and not necessarily those of the NIHR, NHSBT or the Department of Health and Social Care. A complete list of the investigators and contributors to the INTERVAL trial is provided in ref.^16^.

## Authorship

H.V. conducted all the analyses and wrote the paper. P.T., J.B., C.K. and H.McK. generated FC and Sysmex data for the PFC. N.G. conducted the imputation of the PFC genotyping data. J.D. established INTERVAL and provided critical comments on the manuscript. A.M. provided clinical and biological interpretation. J.H. provided *in vitro* data on thrombus formation phenotypes. W.H.O. established the PFC and INTERVAL. K.D. supervised experiments and oversaw analyses. W.J.A. and E.T. supervised the project and wrote the paper jointly.

Conflict-of-interest disclosure: The authors have no competing interests.

## Footnotes

^*^W.J.A. and E.T. jointly supervised this work.

