## Supplemental Figures for "A signature of platelet reactivity in CBC scattergrams reveals genetic predictors of thrombotic disease risk"

Verdier et al. (2023)

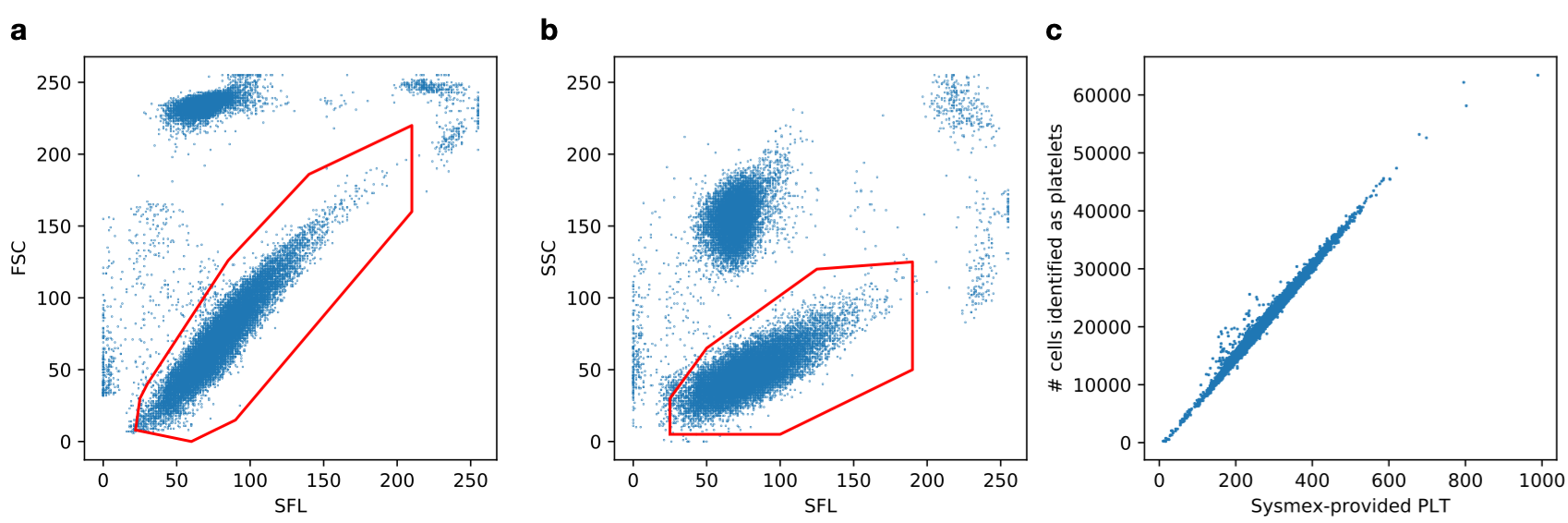
**Supplemental Figure 1 | Gating scheme to identify platelets. a)** The SFL-FSC gate identified using our approach (in red) superimposed on an example of a PLT-F scattergram of a donor from the PFC. **b)** The corresponding SFL-SSC gate, on the same donor. **c)** Scatterplot of PLT against the number of cells identified as platelets by our gating approach (*R*^2^ = 0.99).

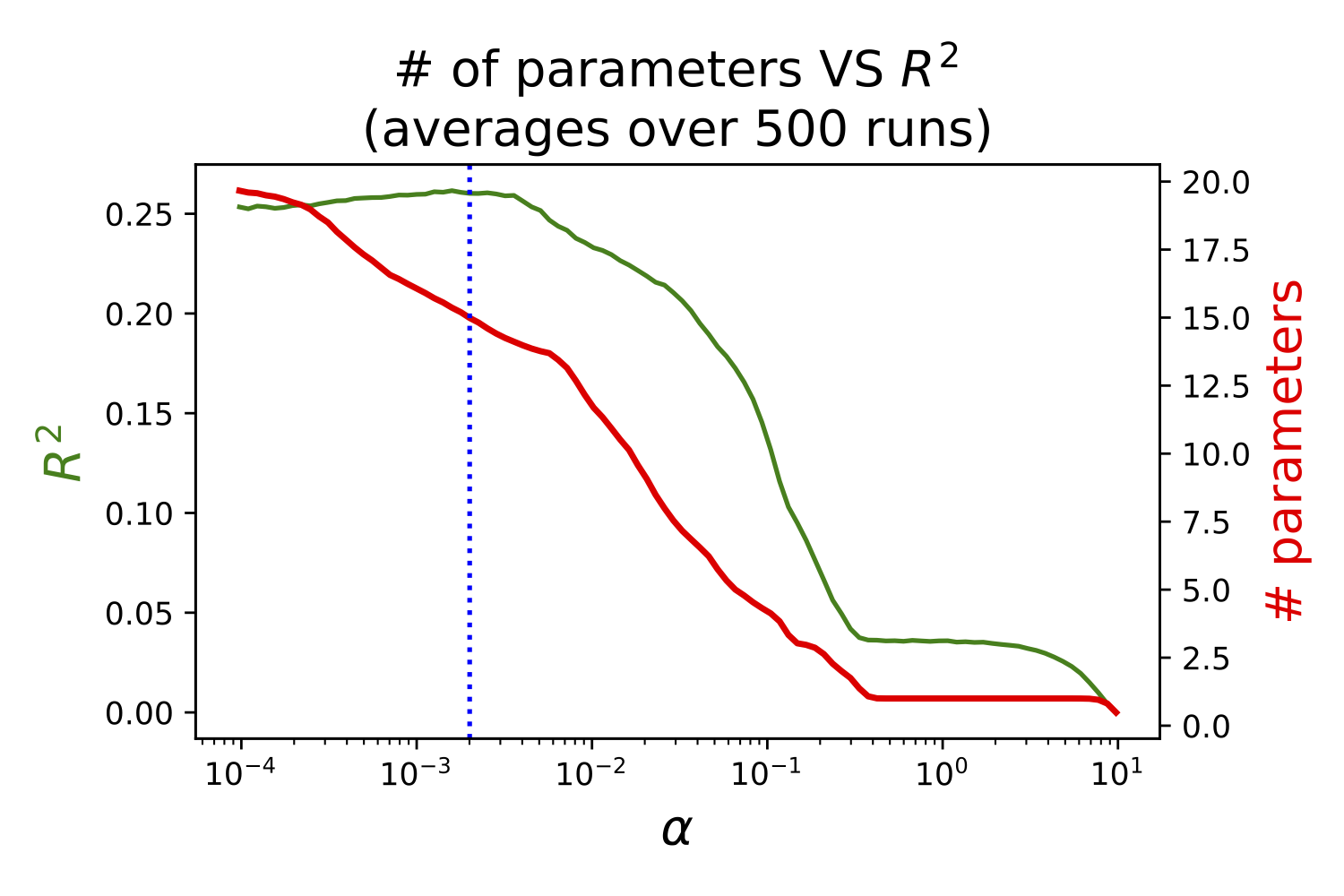

**Supplemental Figure 2 | Setting the hyperparameter of the Lasso regression.** *x*-axis: α, regularisation parameter for the Lasso regression of PR to ADP on the 15 scattergram-derived features and the five standard platelet traits. Left *y*-axis (green line): mean *R*^2^ on test samples, computed on 500 independent 4-fold random splits. Right *y*-axis (red line): mean number of parameters with a non-null coefficient in the regression.

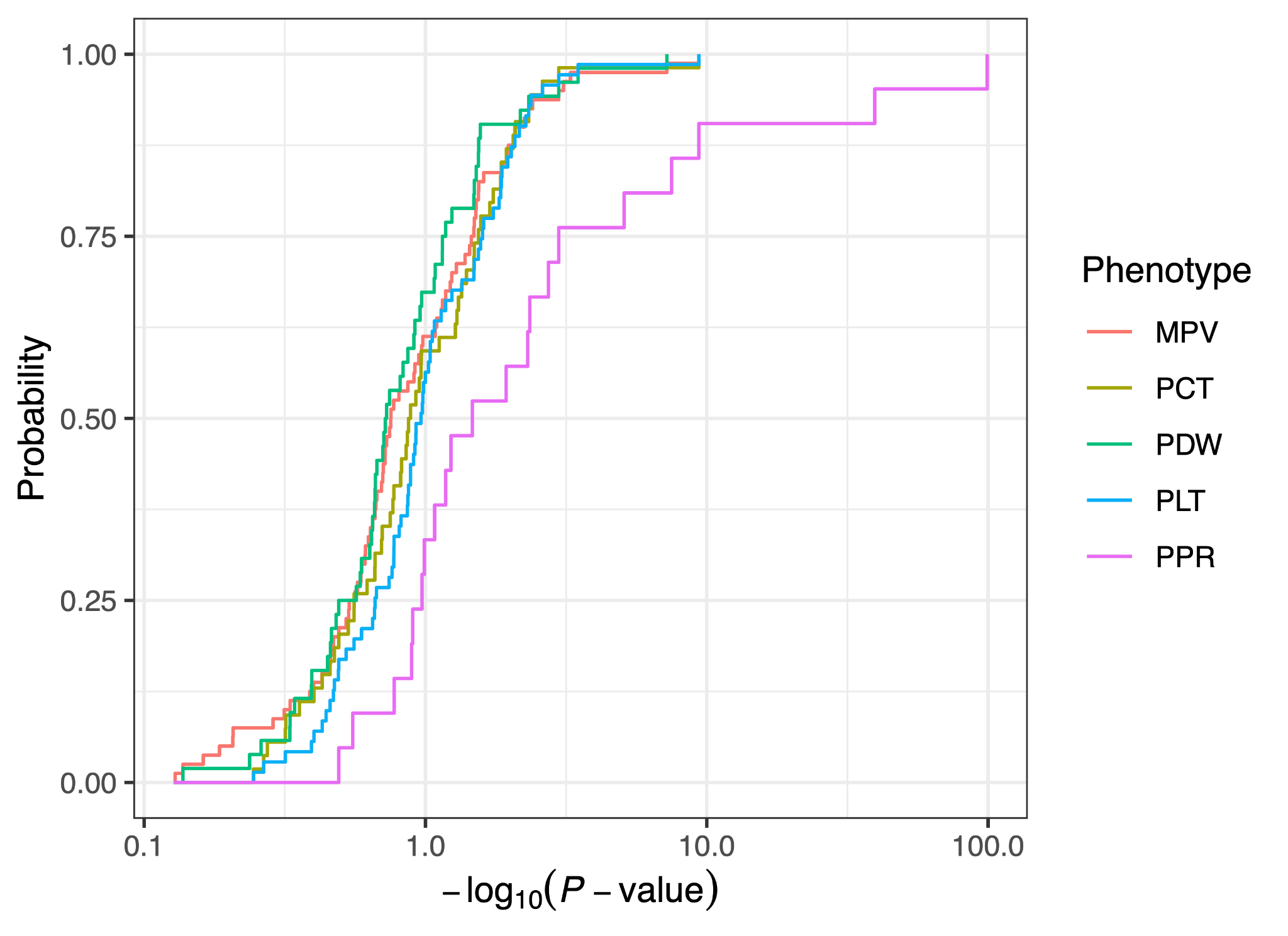

**Supplemental Figure 3 | The distribution of -log_10_ *P*-values for association with PR phenotypes is shifted right amongst variants associated with PPR compared with variants associated with CBC traits in the INTERVAL study.** Empirical cumulative distribution functions of maximum (over the four agonists) -log_10_ *P*-values for genetic association with FC-derived PR in the PFC. Each line corresponds to a different trait and is constructed from the set of variants tagging the genome-wide significant genetic associations in the INTERVAL study.

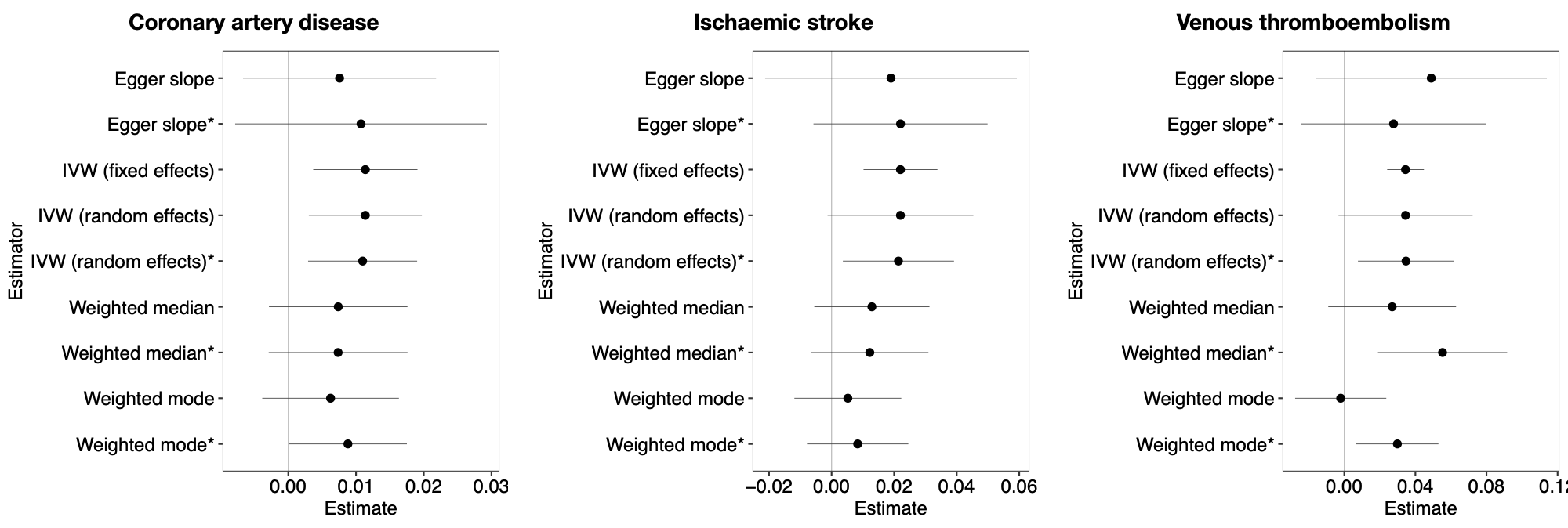

**Supplemental Figure 4 | Results of Mendelian randomization analyses.** Estimates and 95% confidence intervals of the causal effect of general PR on the log odds of a disease event derived from various Mendelian randomization estimators. The analyses denoted with an asterisk include very weak instruments (minimum *P-*value in PFC ≥ 0.05).
