## Supplemental Methods for "A signature of platelet reactivity in CBC scattergrams reveals genetic predictors of thrombotic disease risk"

Verdier *et al.* (2023)

**Measuring flow cytometry-derived platelet reactivity.** Platelet activation in response to different agonists was measured using whole blood flow cytometry (FC) according to a previously published protocol^1,2^. Citrated whole blood was incubated with aspirin and hirudin to block platelet activation mediated through COX-1 and by thrombin, respectively. Treated blood was agonised with each of ADP, CRP-XL, PAR-1 agonist peptide and PAR-4 agonist peptide. The agonist concentrations are given in Supplementary Table 1.4 of Downes et al. (2021)^1^. Platelets were labelled with each of phycoerythrin-conjugated anti-CD62P (P-selectin) monoclonal antibody (Thromb6, Bristol Institute for Transfusion Science, NHSBT Bristol, UK) and polyclonal fluorescein isothiocyanate-labelled anti-fibrinogen antibody (Agilent, F011102-2), a measure of α-granule release and bound fibrinogen respectively. Platelets were identified by FSC and SSC, a measure of size and granularity, and fluorescence levels were compared between agonised and unagonised samples from the same individuals. The corresponding negative control for the P-selectin antibody consisted of an isotype control in the absence of agonist, while the negative control for fibrinogen binding included the addition of EDTA to inhibit the calcium-dependent heterodimerisation of α_IIb_β_3_. Platelet reactivity (PR) was quantified as activation exceeding thresholds set on the isotype and negative controls. All experiments were performed in duplicate. For quality control, we omitted individuals for whom the difference between the logit-transformed percentages of activated platelets observed in the two replicates was greater than the mean plus three times the standard deviation of these differences across the entire cohort. We then averaged the replicate measurements within label, standardised them, and omitted individuals for whom P-selectin secretion and fibrinogen binding were inconsistent using a similar criterion: we discarded those for which the difference between P-selectin and fibrinogen measurements was greater than the average plus three times the standard deviation of the difference. Finally, to obtain a single PR phenotype for a given agonist, we standardised the distribution of P-selectin measurements and the distribution of fibrinogen measurements, averaged each within-individual pair of standardised measurements and standardised the distribution of the averages.

**Isolating platelets in Sysmex XN scattergrams.** A Sysmex XN instrument aspirates 88μl of a biological sample (typically, EDTA-anticoagulated whole blood). Several aliquots are taken from the aspirate in turn, each of which is combined with a proprietary reagent to provide material for a Sysmex measurement channel^3^. Five of these channels generate data using an internal FC, of which, the PLT-F channel is designed to measure properties of platelets. The channel generates a three dimensional scattergram, each point of which corresponds to measurements of a cell in an aliquot of blood treated with a proprietary reagent. The measurements are made by exposing each cell to laser light. The axes of the scattergram represent forward scatter (FSC), side scatter (SSC) and side fluorescence light (SFL). FSC measures the amount of diffracted light, SSC measures the amount of scattered light, and SFL measures the fluorescence of a nucleic acid binding fluorophore. FSC is an approximate measure of a cell's volume, SSC depends on a cell's internal complexity, and SFL captures a cell's nucleic acid content and membrane permeability^4^. Approximately 30,000 cells are measured in a given sample, in two successive phases. In the first phase, approximately 10,000 cells are measured to define a region (a 'gate') on the SFL-FSC plane of the scattergram which contains the highly abundant red blood cells. In the second phase, approximately 20,000 cells residing outside that gate are measured. The instrument computes PLT and IPF from the PLT-F channel's second phase scattergram, presumably by counting the cells lying in a predetermined gate assumed to contain platelets. However, the details of this algorithm are not publicly available. We therefore developed our own gating procedure to identify platelets, using both the SFL-FSC and the SFL-SSC planes. We defined gates in these two planes and considered as platelets all cells lying in both gates (**Supplemental** **Table 2**). We validated our approach by comparing the number of cells lying inside the gates with the corresponding PLT value generated by the instrument in each sample (**Supplemental Figure 1**). The instrument's impedance channel measures the width of each platelet. The data from the PLT-F channel and the impedance channel together determine the five standard platelet traits: PLT, MPV, PCT, PDW and IPF.

**Adjusting for experimental variability in PLT-F scattergrams.** The Sysmex scattergrams for the INTERVAL cohort were collected over two years using two different instruments, both of which differed from the instrument used to phenotype the Cambridge Platelet Function Cohort (PFC). To account for sources of technical variation, we developed a procedure that calibrates the INTERVAL scattergrams to the PFC scattergrams. Our approach adjusts for between-instrument and time-dependent technical variation, allowing predictive models fitted on PFC data to be applied reliably on INTERVAL data. To guard against confounding due to differences in demographic composition between the PFC and INTERVAL cohorts and within INTERVAL over time, we calibrated our adjustment procedure using data from a relatively homogenous population of European ancestry males aged 40–60 years. There were 88 such individuals in the PFC and 7,636 such individuals in the INTERVAL cohort. We extracted the 15 features from the INTERVAL scattergrams, as described above. We then ran a breakpoint detection algorithm based on radial basis functions^5^ on each of the two instrument-specific 15-dimensional time series of the features, identifying sudden shifts in the distributions of these features caused by unknown technical factors. The penalization parameter of the breakpoint detection algorithm was set by trial and error to obtain breakpoints which seemed reasonable for both instruments, by visual inspection, and which led to a minimum separation between two consecutive breakpoints of 150 samples. Within each bin delimited by two adjacent breakpoints, we aggregated the INTERVAL scattergrams. For each bin, we found an affine transformation, defined by a translation and a symmetric matrix, that induced a match between the mean and covariance of the aggregated INTERVAL scattergrams and the aggregated PFC scattergrams. Finally, we applied the transformation computed for each bin to all the available scattergrams generated by the instrument within the time period demarcated by the breakpoints delimiting the bin. This produced adjusted scattergrams for 29,806 INTERVAL participants, from which we extracted the features for prediction using the procedure described above.

**Extracting features from PLT-F scattergrams.** We developed a procedure to extract features from a collection of gated PLT-F second phase scattergrams generated by a given instrument. The features were designed to capture the inter-individual variation between such scattergrams. The procedure relies on loadings obtained by fitting a PCA on a composite set of points comprising 1,000 randomly chosen platelets from each participant in the PFC. The aggregation of an equal number of points from each sample ensures that each individual carries equal weight. These PFC-derived PCA loadings were recorded for the purpose of reducing the dimensionality of scattergrams obtained from any instrument or cohort from three to two. Our procedure extracts 15 features from the two-dimensional 1,000-point cloud obtained after subsampling and dimensionality reduction: the median along each of the two dimensions; the standard deviation along each dimension; the first and last decile along each of the two dimensions; the difference between the last and first quartile along each of the two dimensions; the standard deviation along the second dimension of points in the upper and lower deciles of the first dimension; the logarithm of the ratio of the two standard deviations; the logarithm of the ratio of the two lower deciles; the logarithm of the ratio of the two upper deciles.

**Building a predictive model of FC-derived PR from Sysmex PLT-F scattergrams**. To predict PR in response to a given agonist from the 15 scattergram-derived features (described above) and the five standard platelet traits (PLT, MPV, PCT, PDW and IPF), 20 features in total), we fitted a Lasso regression^6,7^ using data from the 533 participants in the PFC for which both the Sysmex scattergrams and the FC-derived platelet activation measurements were available. We used 100 repetitions of 5-fold cross validation to select the penalty parameter of the Lasso from a grid of 50 logarithmically-uniform spaced values ranging from 1x10^-4^ to 1 in order to maximise the mean (over repetitions and folds) *R^2^* of the regression on the held-out data. In the case PR to ADP, the optimal value of the penalty parameter yielded a mean prediction *R*^2^=0.26. The mean prediction *R^2^* was not overly sensitive to the value of the penalty parameter around the maximum (**Supplemental Figure 2**). Finally, we fitted a linear regression model of the ADP response on the 14 covariates with non-zero coefficients in the cross-validation, using the entire dataset.

**Genome-wide association study of predicted PR in INTERVAL.** We predicted PR in response to ADP in the INTERVAL cohort using the model fitted to the PFC data (**Methods**). The INTERVAL cohort was genotyped using the UK Biobank Affymetrix array. Unmeasured variants were imputed from a combined 1000 Genomes Phase 3-UK10K whole genome sequencing panel. The methods used to QC the measured genotypes and to impute the unmeasured genotypes have been previously described^8^. After discarding variants with a minor allele frequency less than 1% and an imputation INFO score less than 0.4 we sought, we performed a GWAS of the predicted phenotype in the European ancestry subset of INTERVAL(*n*=29,806) using BOLT-LMM^9^ including the following covariates: sex, the score vectors corresponding to the ten leading principal components of within sample genetic variation, and a categorical variable corresponding to whether the scattergram was measured from a blood sample taken at baseline or at 2 or 4 year follow up. We declared variants to be significantly associated if they passed the genome-wide significance threshold of *P*<10^-8^. To identify a parsimonious subset of genetic variants explaining the significant associations, we a) grouped variants by genomic location using complete linkage hierarchical clustering, ensuring that any two variants less than 5Mb apart were assigned to the same cluster^10^, and b) applied a stepwise model selection procedure to each set of variants^8^.

**Imputation of genotypes in the PFC.** The genotypes in the PFC were imputed from a combined 1000 Genomes Phase 3-UK10K whole genome sequencing panel as described in Downes et al. (ref.^1^).

**Regression of FC-derived phenotypes on variant imputed allele counts.** We regressed the four FC-derived PR phenotypes on the imputed allele counts of the 21 predicted PR (PPR)-associated variants (identified in INTERVAL) using data from 1,373 participants in the PFC^1^. Two PPR-associated variants (rs3819288 and rs59001897) were missing from the imputed genotypes in the PFC. To address this, we used the LDproxy tool^11^ to identify the most strongly correlated alternative variant in the reference panel of British in England and Scotland that had been typed in the PFC.

**Replication in Keramarti et al. LTA study.** We downloaded the publicly available summary statistics from the LTA study of Keramati et al.^12^, which are restricted to the set of variants with association test *P*-values < 3×10^-4^ (https://ftp.ncbi.nlm.nih.gov/dbgap/studies/phs001974/analyses/). We identified the variant in summary statistics exhibiting the strongest LD (*r*^2^ in INTERVAL Europeans) with each of the 21 PPR associated variants and the phenotype exhibiting the smallest *P*-value for association with that variant.

**Regression of an *in vitro* thrombus formation phenotype on variant imputed allele counts.** A 48-dimensional *in vitro* thrombus formation phenotype was measured on 87 genotyped participants in the PFC^13^. After standardising each dimension to have a mean of zero and a variance of one, we performed a principal component analysis. We regressed the leading principal component on the imputed variant allele counts corresponding to the 21 PPR-associated variants identified in INTERVAL. We compared the *P*-values and effect sizes identified in the genetic analysis of PPR in INTERVAL and the genetic analysis of the *in vitro* formation phenotype in the PFC. We used proxies for two PPR-associated variants (rs3819288 and rs59001897) as described above.

**Building a genetic score of general PR.** For each of the variants identified by the INTERVAL GWAS (or the corresponding proxy, see above), we obtained the vector of previously published PFC effect sizes with respect to FC-measured PR in response to four agonists^1^. We sought to calibrate the effect sizes of the genetic variants, to put them on a scale measuring a general propensity of platelets to activate (a latent, agonist-independent form of PR). Assuming that each causal variant is involved in one of the four measured activation pathways, we linked each variant to the agonist yielding the smallest *P*-value of association in the PFC and assigned it to that pathway. We then applied a standardisation procedure to calibrate the effect sizes corresponding to each agonist. For each agonist and the corresponding subset of the 21 variants assigned to the pathway of that agonist, we fitted a least squares regression of the effect sizes in INTERVAL on the effect sizes in the PFC, with no intercept term. The estimated regression coefficients represented agonist-specific calibration factors that we used to place the effect sizes from the PFC on approximately the same scale as the effect sizes in INTERVAL. Finally, we computed a polygenic score of general PR as the sum of the linear combinations of genotypes and effect sizes within each of the four agonists.

**Survival analysis in UK Biobank.** We performed Cox regression analyses in UK Biobank to test for associations between the genetic score of PR and health outcomes derived from electronic health records. ICD10 code data are recorded at two levels of granularity in UK Biobank: a coarse level consisting of a three letter code, and a granular level consisting of a three letter code followed by a decimal point and an integer. To harmonise the ICD10 codes, we converted all granular ICD10 codes to the corresponding coarse codes. For any individuals with more than one coarse ICD10 code, we retained only the earliest event. We restricted the association analysis to the 524 (coarse) ICD10 codes that we identified as having at least 1,000 cases among 384,059 European-ancestry participants. For each of 524 outcomes, we performed a Cox proportional-hazards regression accounting for several factors known to play a role in cardiovascular diseases: sex, tobacco use, total cholesterol level, HDL cholesterol level, systolic blood pressure, C-reactive protein blood level and history of diabetes^14^. Tobacco use was accounted for using the number of pack years of smoking divided by the age of the individuals minus fifteen years, so as to obtain a variable indicative of the average tobacco consumption rate over adult life. As some of the continuous covariates may not respect the proportional hazards assumption, we identified quantiles for each covariate (either tertiles or quartiles) and, in conjunction with the binary covariates, we used them to stratify the participants into 864 bins within which base hazard curves were estimated^5^. For each ICD10 code, we included PLT, MPV, PCT and PDW as covariates and obtained *P*-values using a likelihood-ratio test with one degree of freedom, corresponding to the comparison of models with and without our score.

**Mendelian randomisation analyses.** We performed two-sample Mendelian randomisation analyses to estimate the causal effect of general PR (variant effect sizes calibrated as described in **Building a genetic score of general PR** above) on the log odds of disease events for three cardiovascular diseases^15^. We extracted the estimates of the log odds ratios and the corresponding standard errors from the summary statistic files of recent large GWAS of coronary artery disease^16^, stroke^17^ and venous thromboembolism^18^ in European ancestry participants. We excluded rs61751937 because evidence in the literature suggests variation in *SVEP1* expression may be a risk factor for atherosclerosis^19^ (i.e., there may be horizontal pleiotropy). To reduce the effect of weak instrument bias (which in the two-sample MR context is towards the null) we selected the ten remaining variants in **Table 1** with a *P*-value of association (with a PR phenotype) less than 0.05 in the PFC as primary instruments. We meta-analysed the instrument specific ratio estimates for the ten variants using the standard inverse variance weighted (IVW) fixed effects estimator, with second order weights (**Figure 4e–g**). We then performed a series of secondary analyses using both the primary instruments and the complete set of variants (i.e., the 20 variants excluding rs61751937), including very weak instruments. MR Egger random effects regression analyses suggested no evidence of unbalanced horizontal pleiotropy^20^. We also calculated the IVW random effects, weighted median and weighted mode estimates and their corresponding 95% confidence intervals^21,22^.
